# Underdiagnosis of myalgic encephalomyelitis/chronic fatigue syndrome-like illness in a large integrated healthcare system — Kaiser Permanente Northern California, 2022-2023

**DOI:** 10.1101/2024.12.04.24318508

**Authors:** Mariah S. Wood, Nicole Halmer, Jeanne Bertolli, Laura B. Amsden, Joshua R. Nugent, Jin-Mann S. Lin, Gretchen Rothrock, Joelle Nadle, Shua J. Chai, Jamila H. Champsi, James Yang, Elizabeth R. Unger, Jacek Skarbinski, STOP-ME/CFS and COVID-SELECT

## Abstract

**Background:** Surveillance of myalgic encephalomyelitis/chronic fatigue syndrome (ME/CFS), a chronic, debilitating multisystem illness, is challenging because ME/CFS can be under-recognized in healthcare settings.

**Methods:** Using a population-based panel study of 9,820 adult members of Kaiser Permanente Northern California (KPNC), a large, integrated healthcare system, we compared survey-defined ME/CFS-like illness with presence of an ME/CFS diagnosis in the electronic health record (EHR) to evaluate ME/CFS underdiagnosis.

**Results:** Of those with survey-defined ME/CFS-like illness, an estimated 97.8% (95% confidence interval [CI] 97.1%-98.4%) did not have an ME/CFS diagnosis in the EHR. The group without EHR diagnosis was younger, less likely to identify as white non-Hispanic, and more likely to have developed fatigue in the past three years than the EHR diagnosed group. Both diagnosed and undiagnosed ME/CFS-like illness groups had significantly impaired physical, cognitive, and social functioning, and significantly worse mental health and anxiety than those without ME/CFS-like illness.

**Conclusion:** ME/CFS is underdiagnosed in the Kaiser Permanente Northern California healthcare system. Enhanced syndromic surveillance that characterizes patients with ME/CFS who have not been diagnosed has the potential to increase timely recognition of ME/CFS.

## Introduction

Myalgic encephalomyelitis/chronic fatigue syndrome (ME/CFS) is a chronic, debilitating multisystem illness that affects millions of people worldwide (1). The Institute of Medicine (IOM) 2015 diagnostic criteria require a substantial reduction in the ability to engage in pre-illness activity for more than six months, accompanied by fatigue of new or definite onset not resulting from excessive exertion and that is unrelieved by rest, post-exertional malaise, unrefreshing sleep, and either cognitive impairment or orthostatic intolerance (2,3). Given the lack of a diagnostic biomarker, ME/CFS is diagnosed by clinicians based on patient self-report of symptoms that meet the IOM diagnostic criteria after full clinical evaluation to identify other conditions that could contribute to symptoms (4). Prior studies have suggested that most ME/CFS patients are undiagnosed (1), and the number of people with undiagnosed ME/CFS may have recently increased given ME/CFS is a possible outcome of SARS-CoV-2 infection (5). Understanding the extent of ME/CFS underdiagnosis is crucial for mobilizing action to improve identification and subsequent care for those with ME/CFS. We estimate the prevalence of undiagnosed ME/CFS-like illness in a large, integrated healthcare system over a two-year study period, compare demographics, healthcare utilization, and health-related quality of life among undiagnosed and diagnosed persons, and suggest measures that could improve rates of diagnosis.

## Methods

In collaboration with Kaiser Permanente Northern California (KPNC), the California Emerging Infections Program conducts enhanced syndromic surveillance for ME/CFS through the Surveillance to Optimize Protocols for Early Identification and Subgrouping of ME/CFS (STOP ME/CFS) project. STOP ME/CFS involves annual surveys to identify and follow patients with ME/CFS-like illness among KPNC’s 4.5 million members, who are demographically similar to the California population (6). The study was approved by the KPNC Institutional Review Board (IRB # 1692449). Informed written consent was obtained for all participants in this study. This activity was reviewed by the CDC and was conducted consistent with applicable federal law and CDC policy.

A stratified random sample of adult English-speaking KPNC members who had been members for at least one year and who had a valid email address on file were recruited from seven mutually-exclusive strata, including: 1) ME/CFS diagnosis in the electronic health record (EHR); 2) post-COVID conditions diagnosis in the EHR; 3) persons at higher risk for ME/CFS based on an internally-created predictive model; 4) persons who had COVID-19 before July 5, 2021; 5) persons who had COVID-19 between July 5, 2021 and December 14, 2021; 6) persons who had COVID-19 after December 15, 2021 through beginning of recruitment (May 9, 2022); and 7) persons who did not fit into any above strata. (For full sampling strategy see methods section and flow diagram in the prior paper stemming from the STOP ME/CFS project (7)).

To select for people who were more likely to have the relatively rare outcome of ME/CFS, persons were oversampled from strata one to three. In stratum one, 8,061 (98.5%) of 8,182 persons were sampled. In stratum two, 6,057 (98.9%) of 6,126 were sampled. In stratum three, 29,720 (99.1%) of 30,000 were sampled. Within each of strata four through seven, a simple random sample was applied. In stratum four, 11,046 (8.3%) of 133,114 were sampled. In stratum five, 10,389 (18.2%) of 57,127 were sampled. In stratum six, 10,982 (8.6%) of 128,206 were sampled and in stratum seven, 22,250 (0.9%) of 2,382,619 persons were sampled. Sampled persons were invited to participate in an online survey from July 10 to October 17, 2022. Of 98,505 eligible sampled members, 9,825 (10.0%) completed the survey in year one. In year two, all respondents from year one were contacted again from May 1 to November 27, 2023; of those respondents, 6,393 (65.1%) completed the survey again in year two (since recruitment, five respondents requested data removal; the updated number of respondents is 9,820). Participants were offered a $10 gift card each time they completed a survey.

The survey assessed sociodemographic characteristics, IOM ME/CFS diagnostic criteria based on standardized scores on symptom questionnaires, COVID-19 history, and health-related quality of life using established instruments (see footnote in Table 2)). Linked EHR data were used to assess ME/CFS and related diagnoses, COVID-19 history, body mass index (BMI), Charlson comorbidity index score, and outpatient primary care utilization (in-person, video, and telephone visits with a provider in family medicine, internal medicine, or obstetrics/gynecology). Of the respondents, 645 in year one and 316 in year two (not mutually exclusive) met criteria for survey-identified ME/CFS-like illness based on IOM ME/CFS diagnostic criteria (Supplemental Table 1). In total, 798 persons had ME/CFS-like illness in either year one, year two, or both. Persons with survey-identified ME/CFS-like illness were classified as either having undiagnosed ME/CFS if they had no recorded ME/CFS diagnosis in their EHR at any time (identified by either International Classification of Disease [ICD]-9 codes 780.71 or 323.9, or ICD-10 codes G93.32, R53.82, or umbrella codes G93.3) or having diagnosed ME/CFS if a ME/CFS diagnosis was documented in their EHR. The primary outcome of interest was prevalence of an EHR ME/CFS diagnosis among those who qualified for survey-identified ME/CFS-like illness. Secondary outcomes of interest included characteristics and functional status of respondents stratified by presence of an EHR ME/CFS diagnosis.

**Table 1.**
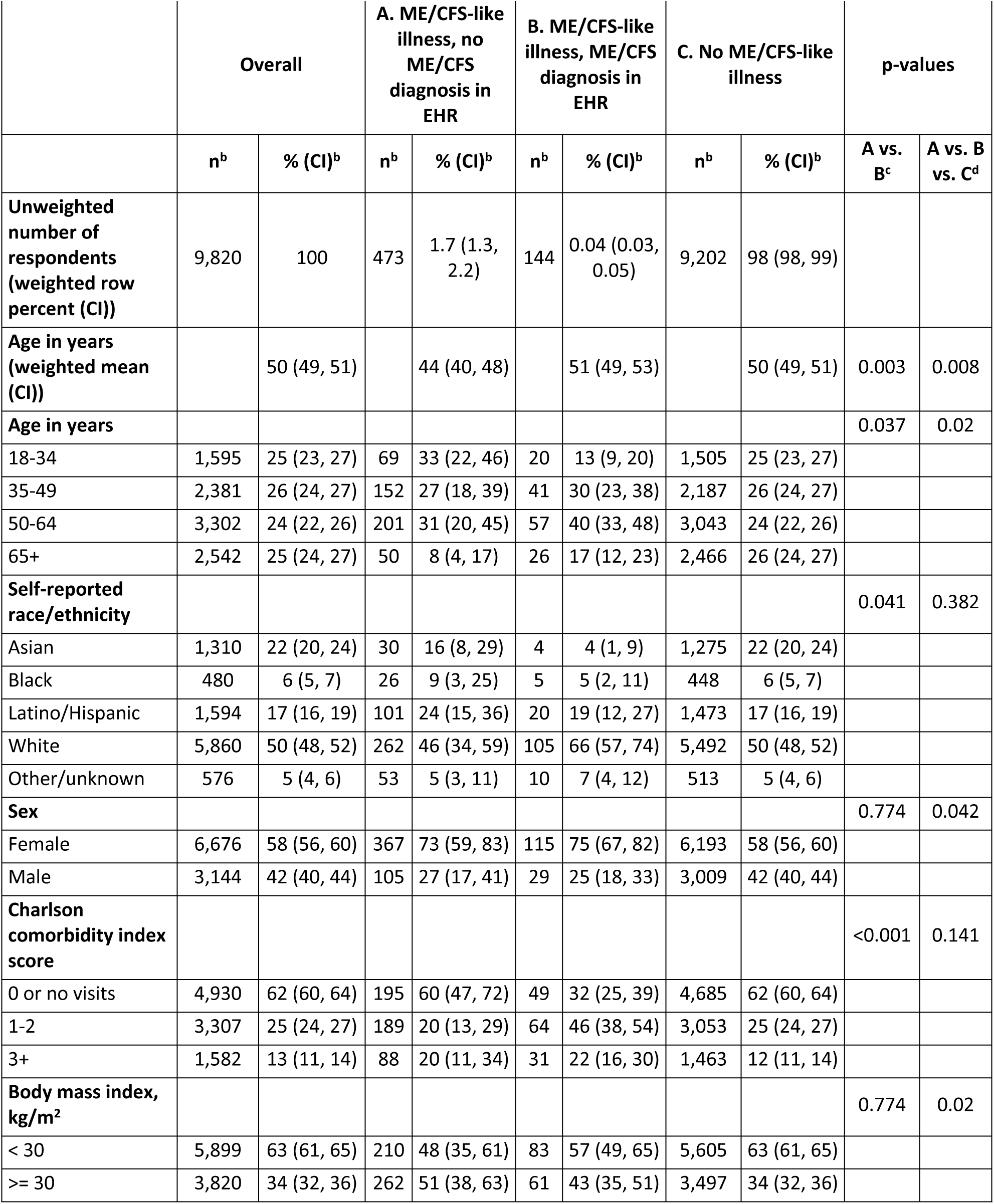
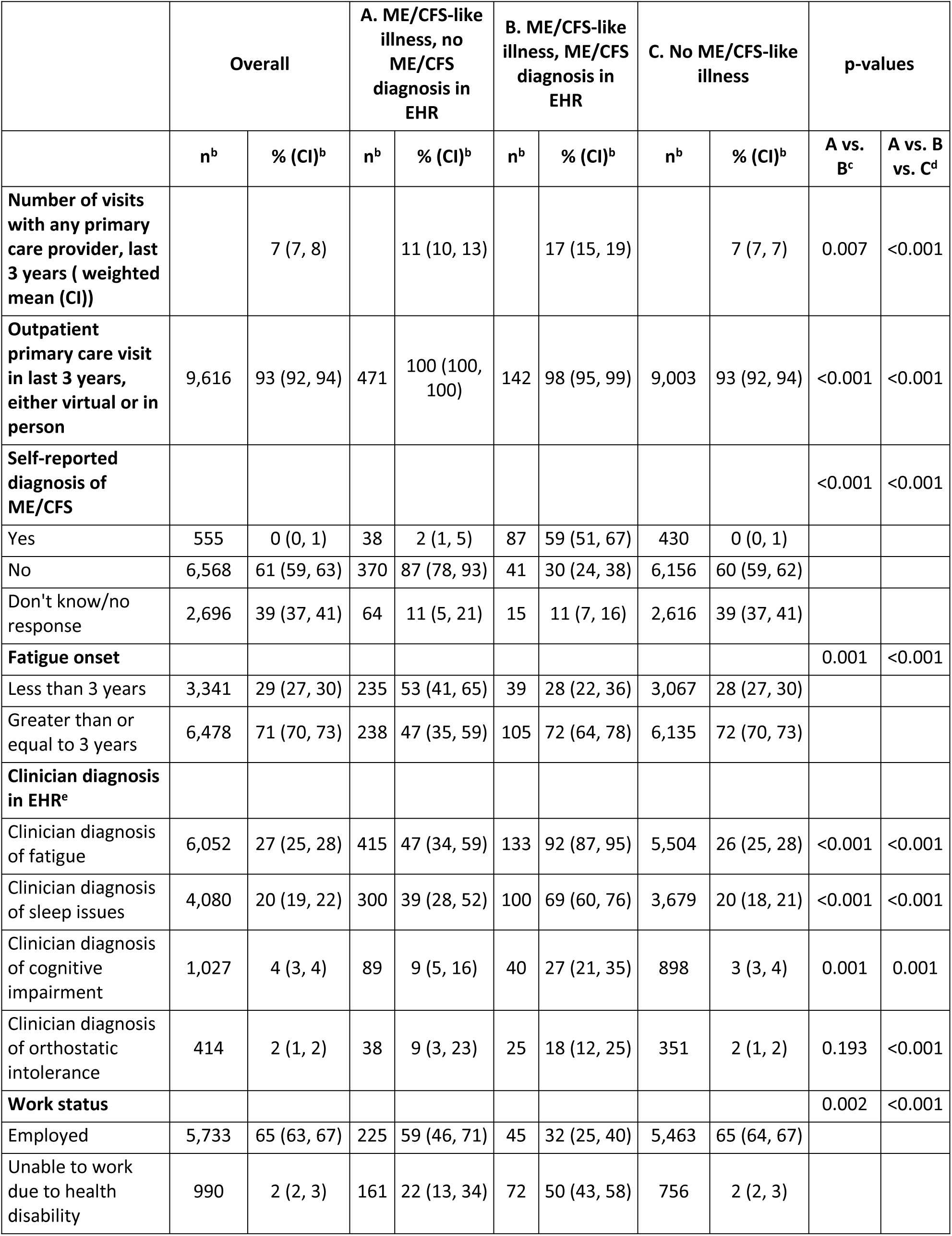
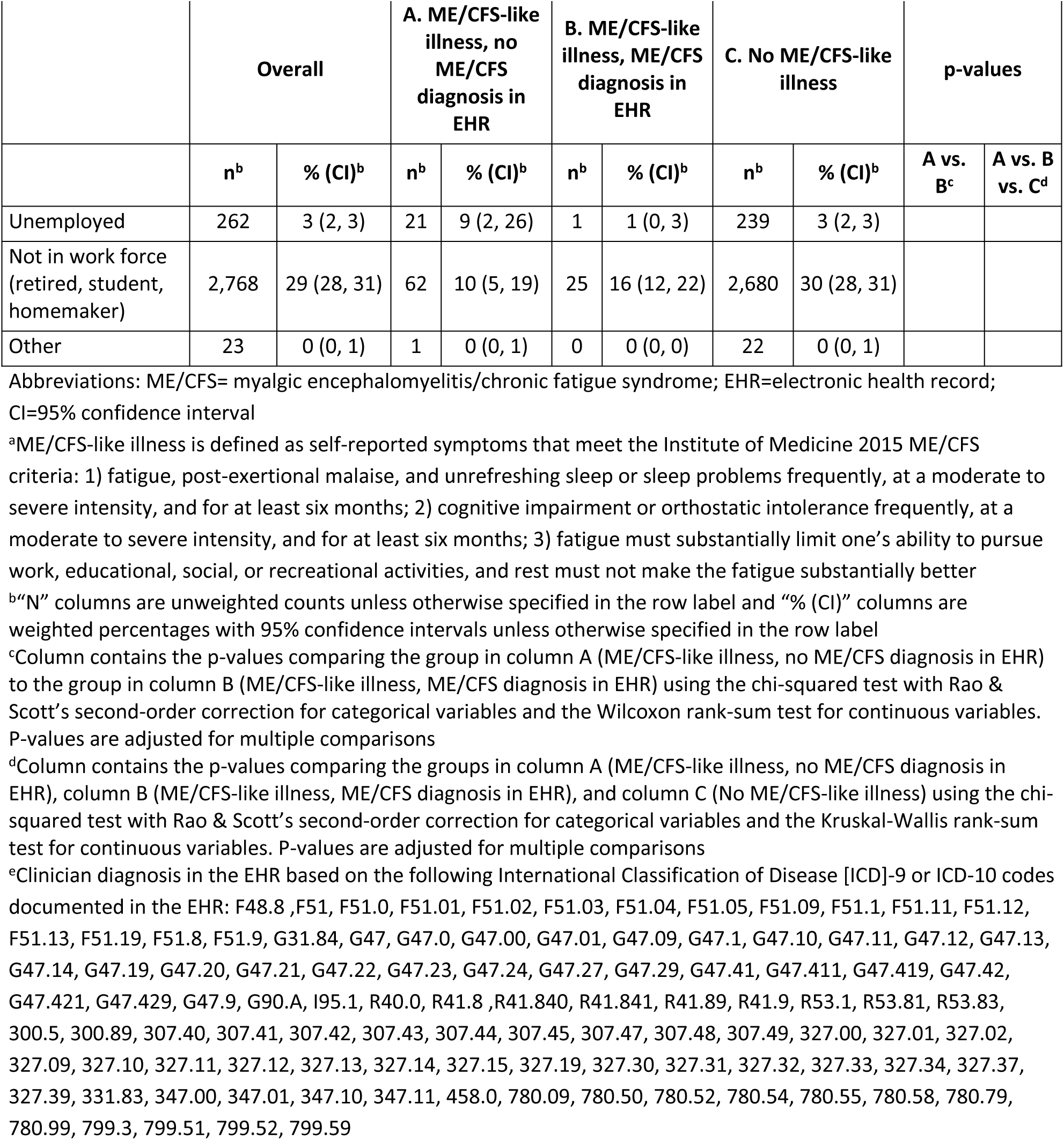
Demographics and characteristics of persons with myalgic encephalomyelitis/chronic fatigue syndrome (ME/CFS)-like illness^a^ on survey, by presence of ME/CFS diagnosis in the electronic health record (EHR)

|  | Overall |  | A. ME/CFS-like illness, no ME/CFS diagnosis in EHR |  | B. ME/CFS-like illness, ME/CFS diagnosis in EHR |  | C. No ME/CFS-like illness |  | p-values |  |
| --- | --- | --- | --- | --- | --- | --- | --- | --- | --- | --- |
|  | n<sup>b</sup> | % (CI)<sup>b</sup> | n<sup>b</sup> | % (CI)<sup>b</sup> | n<sup>b</sup> | % (CI)<sup>b</sup> | n<sup>b</sup> | % (CI)<sup>b</sup> | A vs. B<sup>c</sup> | A vs. B vs. C<sup>d</sup> |
| **Unweighted number of respondents (weighted row percent (CI))** | 9,820 | 100 | 473 | 1.7 (1.3, 2.2) | 144 | 0.04 (0.03, 0.05) | 9,202 | 98 (98, 99) |  |  |
| **Age in years (weighted mean (CI))** |  | 50 (49, 51) |  | 44 (40, 48) |  | 51 (49, 53) |  | 50 (49, 51) | 0.003 | 0.008 |
| **Age in years** |  |  |  |  |  |  |  |  | 0.037 | 0.02 |
| 18-34 | 1,595 | 25 (23, 27) | 69 | 33 (22, 46) | 20 | 13 (9, 20) | 1,505 | 25 (23, 27) |  |  |
| 35-49 | 2,381 | 26 (24, 27) | 152 | 27 (18, 39) | 41 | 30 (23, 38) | 2,187 | 26 (24, 27) |  |  |
| 50-64 | 3,302 | 24 (22, 26) | 201 | 31 (20, 45) | 57 | 40 (33, 48) | 3,043 | 24 (22, 26) |  |  |
| 65+ | 2,542 | 25 (24, 27) | 50 | 8 (4, 17) | 26 | 17 (12, 23) | 2,466 | 26 (24, 27) |  |  |
| **Self-reported race/ethnicity** |  |  |  |  |  |  |  |  | 0.041 | 0.382 |
| Asian | 1,310 | 22 (20, 24) | 30 | 16 (8, 29) | 4 | 4 (1, 9) | 1,275 | 22 (20, 24) |  |  |
| Black | 480 | 6 (5, 7) | 26 | 9 (3, 25) | 5 | 5 (2, 11) | 448 | 6 (5, 7) |  |  |
| Latino/Hispanic | 1,594 | 17 (16, 19) | 101 | 24 (15, 36) | 20 | 19 (12, 27) | 1,473 | 17 (16, 19) |  |  |
| White | 5,860 | 50 (48, 52) | 262 | 46 (34, 59) | 105 | 66 (57, 74) | 5,492 | 50 (48, 52) |  |  |
| Other/unknown | 576 | 5 (4, 6) | 53 | 5 (3, 11) | 10 | 7 (4, 12) | 513 | 5 (4, 6) |  |  |
| **Sex** |  |  |  |  |  |  |  |  | 0.774 | 0.042 |
| Female | 6,676 | 58 (56, 60) | 367 | 73 (59, 83) | 115 | 75 (67, 82) | 6,193 | 58 (56, 60) |  |  |
| Male | 3,144 | 42 (40, 44) | 105 | 27 (17, 41) | 29 | 25 (18, 33) | 3,009 | 42 (40, 44) |  |  |
| **Charlson comorbidity index score** |  |  |  |  |  |  |  |  | <0.001 | 0.141 |
| 0 or no visits | 4,930 | 62 (60, 64) | 195 | 60 (47, 72) | 49 | 32 (25, 39) | 4,685 | 62 (60, 64) |  |  |
| 1-2 | 3,307 | 25 (24, 27) | 189 | 20 (13, 29) | 64 | 46 (38, 54) | 3,053 | 25 (24, 27) |  |  |
| 3+ | 1,582 | 13 (11, 14) | 88 | 20 (11, 34) | 31 | 22 (16, 30) | 1,463 | 12 (11, 14) |  |  |
| **Body mass index, kg/m<sup>2</sup>** |  |  |  |  |  |  |  |  | 0.774 | 0.02 |
| < 30 | 5,899 | 63 (61, 65) | 210 | 48 (35, 61) | 83 | 57 (49, 65) | 5,605 | 63 (61, 65) |  |  |
| >= 30 | 3,820 | 34 (32, 36) | 262 | 51 (38, 63) | 61 | 43 (35, 51) | 3,497 | 34 (32, 36) |  |  |

|  | Overall |  | A. ME/CFS-like illness, no ME/CFS diagnosis in EHR |  | B. ME/CFS-like illness, ME/CFS diagnosis in EHR |  | C. No ME/CFS-like illness |  | p-values |  |
| --- | --- | --- | --- | --- | --- | --- | --- | --- | --- | --- |
|  | n<sup>b</sup> | % (CI)<sup>b</sup> | n<sup>b</sup> | % (CI)<sup>b</sup> | n<sup>b</sup> | % (CI)<sup>b</sup> | n<sup>b</sup> | % (CI)<sup>b</sup> | A vs. B<sup>c</sup> | A vs. B vs. C<sup>d</sup> |
| **Number of visits with any primary care provider, last 3 years ( weighted mean (CI))** |  | 7 (7, 8) |  | 11 (10, 13) |  | 17 (15, 19) |  | 7 (7, 7) | 0.007 | <0.001 |
| **Outpatient primary care visit in last 3 years, either virtual or in person** | 9,616 | 93 (92, 94) | 471 | 100 (100, 100) | 142 | 98 (95, 99) | 9,003 | 93 (92, 94) | <0.001 | <0.001 |
| **Self-reported diagnosis of ME/CFS** |  |  |  |  |  |  |  |  | <0.001 | <0.001 |
| Yes | 555 | 0 (0, 1) | 38 | 2 (1, 5) | 87 | 59 (51, 67) | 430 | 0 (0, 1) |  |  |
| No | 6,568 | 61 (59, 63) | 370 | 87 (78, 93) | 41 | 30 (24, 38) | 6,156 | 60 (59, 62) |  |  |
| Don't know/no response | 2,696 | 39 (37, 41) | 64 | 11 (5, 21) | 15 | 11 (7, 16) | 2,616 | 39 (37, 41) |  |  |
| **Fatigue onset** |  |  |  |  |  |  |  |  | 0.001 | <0.001 |
| Less than 3 years | 3,341 | 29 (27, 30) | 235 | 53 (41, 65) | 39 | 28 (22, 36) | 3,067 | 28 (27, 30) |  |  |
| Greater than or equal to 3 years | 6,478 | 71 (70, 73) | 238 | 47 (35, 59) | 105 | 72 (64, 78) | 6,135 | 72 (70, 73) |  |  |
| **Clinician diagnosis in EHR<sup>e</sup>** |  |  |  |  |  |  |  |  |  |  |
| Clinician diagnosis of fatigue | 6,052 | 27 (25, 28) | 415 | 47 (34, 59) | 133 | 92 (87, 95) | 5,504 | 26 (25, 28) | <0.001 | <0.001 |
| Clinician diagnosis of sleep issues | 4,080 | 20 (19, 22) | 300 | 39 (28, 52) | 100 | 69 (60, 76) | 3,679 | 20 (18, 21) | <0.001 | <0.001 |
| Clinician diagnosis of cognitive impairment | 1,027 | 4 (3, 4) | 89 | 9 (5, 16) | 40 | 27 (21, 35) | 898 | 3 (3, 4) | 0.001 | 0.001 |
| Clinician diagnosis of orthostatic intolerance | 414 | 2 (1, 2) | 38 | 9 (3, 23) | 25 | 18 (12, 25) | 351 | 2 (1, 2) | 0.193 | <0.001 |
| **Work status** |  |  |  |  |  |  |  |  | 0.002 | <0.001 |
| Employed | 5,733 | 65 (63, 67) | 225 | 59 (46, 71) | 45 | 32 (25, 40) | 5,463 | 65 (64, 67) |  |  |
| Unable to work due to health disability | 990 | 2 (2, 3) | 161 | 22 (13, 34) | 72 | 50 (43, 58) | 756 | 2 (2, 3) |  |  |

|  | Overall |  | A. ME/CFS-like illness, no ME/CFS diagnosis in EHR |  | B. ME/CFS-like illness, ME/CFS diagnosis in EHR |  | C. No ME/CFS-like illness |  | p-values |  |
| --- | --- | --- | --- | --- | --- | --- | --- | --- | --- | --- |
|  | n<sup>b</sup> | % (CI)<sup>b</sup> | n<sup>b</sup> | % (CI)<sup>b</sup> | n<sup>b</sup> | % (CI)<sup>b</sup> | n<sup>b</sup> | % (CI)<sup>b</sup> | A vs. B<sup>c</sup> | A vs. B vs. C<sup>d</sup> |
| Unemployed | 262 | 3 (2, 3) | 21 | 9 (2, 26) | 1 | 1 (0, 3) | 239 | 3 (2, 3) |  |  |
| Not in work force (retired, student, homemaker) | 2,768 | 29 (28, 31) | 62 | 10 (5, 19) | 25 | 16 (12, 22) | 2,680 | 30 (28, 31) |  |  |
| Other | 23 | 0 (0, 1) | 1 | 0 (0, 1) | 0 | 0 (0, 0) | 22 | 0 (0, 1) |  |  |

Abbreviations: ME/CFS= myalgic encephalomyelitis/chronic fatigue syndrome; EHR=electronic health record; CI=95% confidence interval
<sup>a</sup>ME/CFS-like illness is defined as self-reported symptoms that meet the Institute of Medicine 2015 ME/CFS criteria: 1) fatigue, post-exertional malaise, and unrefreshing sleep or sleep problems frequently, at a moderate to severe intensity, and for at least six months; 2) cognitive impairment or orthostatic intolerance frequently, at a moderate to severe intensity, and for at least six months; 3) fatigue must substantially limit one's ability to pursue work, educational, social, or recreational activities, and rest must not make the fatigue substantially better
<sup>b</sup>"N" columns are unweighted counts unless otherwise specified in the row label and "% (CI)" columns are weighted percentages with 95% confidence intervals unless otherwise specified in the row label
<sup>c</sup>Column contains the p-values comparing the group in column A (ME/CFS-like illness, no ME/CFS diagnosis in EHR) to the group in column B (ME/CFS-like illness, ME/CFS diagnosis in EHR) using the chi-squared test with Rao & Scott's second-order correction for categorical variables and the Wilcoxon rank-sum test for continuous variables. P-values are adjusted for multiple comparisons
<sup>d</sup>Column contains the p-values comparing the groups in column A (ME/CFS-like illness, no ME/CFS diagnosis in EHR), column B (ME/CFS-like illness, ME/CFS diagnosis in EHR), and column C (No ME/CFS-like illness) using the chi-squared test with Rao & Scott's second-order correction for categorical variables and the Kruskal-Wallis rank-sum test for continuous variables. P-values are adjusted for multiple comparisons
<sup>e</sup>Clinician diagnosis in the EHR based on the following International Classification of Disease [ICD]-9 or ICD-10 codes documented in the EHR: F48.8, F51, F51.0, F51.01, F51.02, F51.03, F51.04, F51.05, F51.09, F51.1, F51.11, F51.12, F51.13, F51.19, F51.8, F51.9, G31.84, G47, G47.0, G47.00, G47.01, G47.09, G47.1, G47.10, G47.11, G47.12, G47.13, G47.14, G47.19, G47.20, G47.21, G47.22, G47.23, G47.24, G47.27, G47.29, G47.41, G47.411, G47.419, G47.42, G47.421, G47.429, G47.9, G90.A, I95.1, R40.0, R41.8, R41.840, R41.841, R41.89, R41.9, R53.1, R53.81, R53.83, 300.5, 300.89, 307.40, 307.41, 307.42, 307.43, 307.44, 307.45, 307.47, 307.48, 307.49, 327.00, 327.01, 327.02, 327.09, 327.10, 327.11, 327.12, 327.13, 327.14, 327.15, 327.19, 327.30, 327.31, 327.32, 327.33, 327.34, 327.37, 327.39, 331.83, 347.00, 347.01, 347.10, 347.11, 458.0, 780.09, 780.50, 780.52, 780.54, 780.55, 780.58, 780.79, 780.99, 799.3, 799.51, 799.52, 799.59

**Table 2.** Well-being scores of persons with myalgic encephalomyelitis/chronic fatigue syndrome (ME/CFS)-like illness^a^ on survey, by presence of ME/CFS diagnosis in the electronic health record (EHR)

|  |  |  |  |  |  |  |  |  |
| --- | --- | --- | --- | --- | --- | --- | --- | --- |
|  | **A. ME/CFS-like illness, no ME/CFS diagnosis in EHR** |  | **B. ME/CFS-like illness, ME/CFS diagnosis in EHR** |  | **C. No ME/CFS-like illness** |  | **p-values** |  |
| **Characteristic<sup>b</sup>** | **mean<sup>c</sup>** | **CI<sup>c</sup>** | **mean<sup>c</sup>** | **CI<sup>c</sup>** | **mean<sup>c</sup>** | **CI<sup>c</sup>** | **A vs. B<sup>d</sup>** | **A vs. B vs. C<sup>e</sup>** |
| **SF-36** |  |  |  |  |  |  |  |  |
| Physical functioning (T-score) | 38 | (34, 42) | 31 | (29, 32) | 50 | (49, 50) | 0.001 | <0.001 |
| Role-physical (T-score) | 33 | (30, 37) | 26 | (24, 27) | 50 | (50, 50) | <0.001 | <0.001 |
| Bodily pain (T-score) | 35 | (33, 38) | 35 | (33, 36) | 51 | (51, 51) | 0.6 | <0.001 |
| Vitality (T-score) | 30 | (28, 32) | 28 | (27, 29) | 49 | (48, 49) | 0.033 | <0.001 |
| General health (T-score) | 34 | (31, 37) | 28 | (27, 30) | 47 | (47, 47) | <0.001 | <0.001 |
| Social functioning (T-score) | 29 | (26, 32) | 25 | (24, 26) | 47 | (46, 47) | 0.049 | <0.001 |
| Role-emotional (T-score) | 30 | (27, 33) | 29 | (27, 32) | 46 | (46, 47) | 0.789 | <0.001 |
| Mental health (T-score) | 30 | (26, 33) | 34 | (32, 36) | 47 | (47, 48) | 0.033 | <0.001 |
| **PHQ-8 depression score (0-24)** | 16 | (14, 17) | 15 | (14, 16) | 6 | (5, 6) | 0.4 | <0.001 |
| **GAD-2 anxiety score (0-6)** | 4 | (3, 4) | 3 | (3, 3) | 1 | (1, 1) | 0.028 | <0.001 |
| **PROMIS cognitive function (T-score)** | 36 | (34, 38) | 32 | (31, 33) | 52 | (51, 52) | 0.003 | <0.001 |
| **OGS Orthostatic intolerance (0-25)** | 5 | (3, 6) | 5 | (4, 7) | 0 | (0, 0) | 0.4 | <0.001 |

Abbreviations: ME/CFS= myalgic encephalomyelitis/chronic fatigue syndrome; EHR=electronic health record; CI=95% confidence interval
<sup>a</sup>ME/CFS-like illness is defined as self-reported symptoms that meet the Institute of Medicine 2015 ME/CFS criteria: 1) fatigue, post-exertional malaise, and unrefreshing sleep or sleep problems frequently, at a moderate to severe intensity, and for at least six months; 2) cognitive impairment or orthostatic intolerance frequently, at a moderate to severe intensity, and for at least six months; 3) fatigue must substantially limit one's ability to pursue work, educational, social, or recreational activities, and rest must not make the fatigue substantially better
<sup>b</sup>36-Item Short Form Health Survey [SF-36], higher score indicates better functioning; Patient Health Questionnaire Depression scale [PHQ-8], higher score indicates higher levels of depression; Generalized Anxiety Disorder scale [GAD-2], higher score indicates higher levels of anxiety; Patient-Reported Outcomes Measurement Information System cognition instrument Adult v2.0 - Cognitive Function 4a [PROMIS cognition], higher score indicates better cognition; Orthostatic Grading Scale [OGS], higher score indicates more orthostatic intolerance
<sup>c</sup>"Mean" columns are weighted means, "CI" columns are weighted 95% confidence intervals
<sup>d</sup>Column contains the p-values comparing the group in column A (ME/CFS-like illness, no ME/CFS diagnosis in EHR) to the group in column B (ME/CFS-like illness, ME/CFS diagnosis in EHR) using the Wilcoxon rank-sum test. P-values are adjusted for multiple comparisons
<sup>e</sup>Column contains the p-values comparing the groups in column A (ME/CFS-like illness, no ME/CFS diagnosis in EHR), column B (ME/CFS-like illness, ME/CFS diagnosis in EHR), and column C (No ME/CFS-like illness) using the Kruskal-Wallis rank-sum test. P-values are adjusted for multiple comparisons

Sampling weights were calculated equal to the inverse of the probability of sample selection. Non-response weights were estimated using Super Learner, an ensemble machine learning method that uses a weighted combination of candidate algorithms to optimize predictive performance via cross-validation (8). We modeled the probability of survey response within each stratum, adjusting for current age, race and ethnicity, sex, Charlson comorbidity index (CCI) score within the last year (9,10), and body mass index (BMI) within the last two years and generated non-response weights as the inverse of the predicted probability of response. Sampling strata four to seven had higher maximum non-response weights and were trimmed at the 99^th^ percentile. Sampling and non-response weights were multiplied together to determine final survey weights. The weighted sample was compared to the original eligible population using standardized mean differences to ensure the demographic distribution between sample and eligible population was not substantially different (Supplemental Table 3 in the prior paper stemming from the STOP ME/CFS project (7)). For those who had repeated observations (responded both years), final survey weights were divided by two, and half of each weight was applied to each year. All estimates are for the two-year period and include sampling weights and non-response weights as well as adjustment for repeated observations on participants who responded in both years one and two. SF-36 scores and PROMIS cognitive function scores were converted into T-scores (11–13). Associations between the groups were tested using the Wilcoxon rank-sum test for complex survey samples and chi-square test with Rao & Scott’s second-order correction; p-values were adjusted for multiple comparisons (14). Non-response weights were generated using the SuperLearner package (15), weighted, repeated-measures survey data were generated using the survey package (16), and tables of weighted data and p-values were generated using the gtsummary package (17).

## Results

Using a weighted survey of 9,820 adult Kaiser Permanente members over two years, we estimated that 1.8% (95% confidence interval [CI] 1.3%-2.2%) of adult members had survey-identified ME/CFS-like illness during the two-year study period. Among those with survey-identified ME/CFS-like illness, an estimated 97.8% (CI 97.1%-98.4%) did not have any ME/CFS diagnosis in their EHR, with only 2.2% (CI 1.6%-2.9%) having a ME/CFS diagnosis in their health record. Those without an EHR ME/CFS diagnosis were younger (mean age 44 vs. 51) and less likely to identify as White, non-Hispanic (46% vs. 66%) compared to those with an EHR ME/CFS diagnosis (Table 1). Among all those with survey-identified ME/CFS-like illness, the average number of primary care visits over the past three years was 11 (median 9, range 1 to 135); 100% of those without an EHR diagnosis and 98% of those with an EHR diagnosis had at least one virtual or in-person primary care visit in the last three years. Among all those with ME/CFS-like illness, persons without an EHR ME/CFS diagnosis were significantly more likely to report fatigue onset less than three years prior to survey date (53% vs. 28%) than persons with an EHR ME/CFS diagnosis. Moreover, despite reporting these symptoms on the survey, persons with ME/CFS-like illness and without an EHR ME/CFS diagnosis were significantly less likely to have any diagnosis in their EHR of fatigue (47% vs. 92%), sleep issues (39% vs. 69%), or cognitive impairment (9% vs. 27%) compared to persons with ME/CFS-like illness and an EHR ME/CFS diagnosis.

Both groups with survey-identified ME/CFS-like illness, either diagnosed or undiagnosed, reported significantly lower quality of life than a group of KPNC members without ME/CFS-like illness (Table 2). The mean SF-36 T-score was 32 for those with ME/CFS-like illness without an EHR diagnosis, 30 for those with ME/CFS-like illness and an EHR ME/CFS diagnosis, and 48 for those without ME/CFS-like illness; an SF-36 score of 50, with a standard deviation of 10, is the United States population norm (13). Scores on instruments assessing depression, anxiety, cognitive function, and orthostatic symptoms also indicated significant differences for both groups with ME/CFS-like illness compared to the general population. Generally, persons with ME/CFS-like illness and an EHR diagnosis had lower well-being scores than those with ME/CFS-like illness without an EHR diagnosis; however, persons without an EHR ME/CFS diagnosis had a significantly lower SF-36 mental health T-score (30 vs. 34) and significantly higher anxiety score on GAD-2 (worse anxiety) (4 vs. 3) than persons with an EHR ME/CFS diagnosis.

## Discussion

In an integrated healthcare system with 2.7 million adult members, an estimated 1.8% had symptoms consistent with ME/CFS-like illness during a two-year period. Of those with ME/CFS-like illness, 97.8% had no documentation of an ME/CFS diagnosis, despite a high level of primary care utilization; 53% of those with ME/CFS-like illness and no EHR diagnosis reported fatigue began less than three years prior. Surveying medical records alone would not have uncovered this younger, more racially diverse group with recent-onset ME/CFS-like illness. These findings suggest that many people with ME/CFS are not recognized, particularly those with recent onset. Earlier diagnosis of ME/CFS by clinicians might facilitate improvement in outcomes and prevention of disease progression for this group (18). Persons with ME/CFS-like illness without an EHR diagnosis had worse anxiety than persons with ME/CFS-like illness and an EHR diagnosis, suggesting a possible effect of unrecognized illness on mental health among people with ME/CFS symptoms.

This report documents potential underdiagnosis of ME/CFS by healthcare providers, even in an integrated healthcare system with accessible primary care services. Numerous factors may contribute to the high level of underdiagnosis of ME/CFS, including the lack of a diagnostic biomarker, a low level of dissemination of standardized screening and assessment tools, limited uptake of clinician training on ME/CFS, a low-level of patient awareness of ME/CFS, misconceptions among clinicians that the illness is psychogenic in nature or does not exist, and the fact that ME/CFS symptoms can overlap with other illnesses (1,19). The symptoms of the undiagnosed group may provide insight into screening for and monitoring ME/CFS earlier in the illness course.

These findings indicate the importance of enhanced symptom-based surveillance as an initial step in recognizing undiagnosed ME/CFS, as those with undiagnosed ME/CFS-like illness would not have been identified without this project’s symptom-based surveillance efforts. Enhanced syndromic surveillance has the potential to increase timely recognition of ME/CFS. Public communication campaigns could also promote earlier recognition of ME/CFS symptoms, which could lead to earlier care-seeking and care provision. One avenue for public communication is via interactive web-based training for primary healthcare providers and patients. Web-based training could provide resources for managing this complex illness but has had limited implementation to date. Public health detailing, which involves educational visits by public health officials to providers, might also provide support to clinicians, as it has for other chronic conditions (20).

The findings in this report are subject to limitations. First, we assessed ME/CFS-like illness using established IOM ME/CFS criteria based on self-reported symptom frequency, severity, and duration, but did not conduct in-person clinical examinations and interviews to establish a definitive clinical ME/CFS diagnosis; some persons classified as having survey-identified ME/CFS in this study might have an alternative diagnosis or condition that explains their symptoms, which might have inflated our estimate of persons with ME/CFS-like illness. Similarly, we used ICD-9/10 codes to identify persons who had an ME/CFS diagnosis documented in their EHR; ICD-9/10 coding has limitations as a surveillance tool due to the potential for misclassification. Second, our overall survey response rate was 10%; though results were weighted to be representative of the eligible KPNC adult population (see Supplemental Table 3 in prior paper (7)), lower response rates limit the generalizability of results. Third, we only included persons with English listed as their preferred language; 9% of KPNC members prefer a language other than English and were not included in this study.

The results suggest that underdiagnosis of ME/CFS is common despite high levels of care utilization in the Kaiser Permanente Northern California integrated healthcare system. Several strategies could be implemented to attempt to address underdiagnosis, including continued syndromic surveillance, web-based training for providers and patients, and public health detailing. Improved ascertainment of ME/CFS by the medical system is critical for patient care and well-being and has the potential to decrease patient morbidity from ME/CFS through earlier recognition and intervention.

## Data Availability

All available data is presented in the manuscript, tables, figures and supplementary materials in this manuscript. Sharing of additional de-identified data from this study is restricted by the Health Insurance Portability and Accountability Act of 1996 (HIPAA) as a portion of the data is abstracted from electronic health records as part of routine clinical care.

## Acknowledgements

All authors contributed to conceptual ideas and contributed to methodology. MSW, JSL, ERU, JS contributed to operationalization of case-definition and survey instrument scoring; MSW, JRN, JS contributed to formal analysis; MSW, JS contributed to writing and original draft preparation; all authors contributed to writing, reviewing, and editing the manuscript; JS, JB, GR supervised the work. The funders contributed to the study design, decision to publish, and preparation of the manuscript. They did not contribute to the data collection or formal analysis.

The authors are grateful to all Kaiser Permanente members without whom this study would not have been possible.

---

**Supplemental Table 1.**
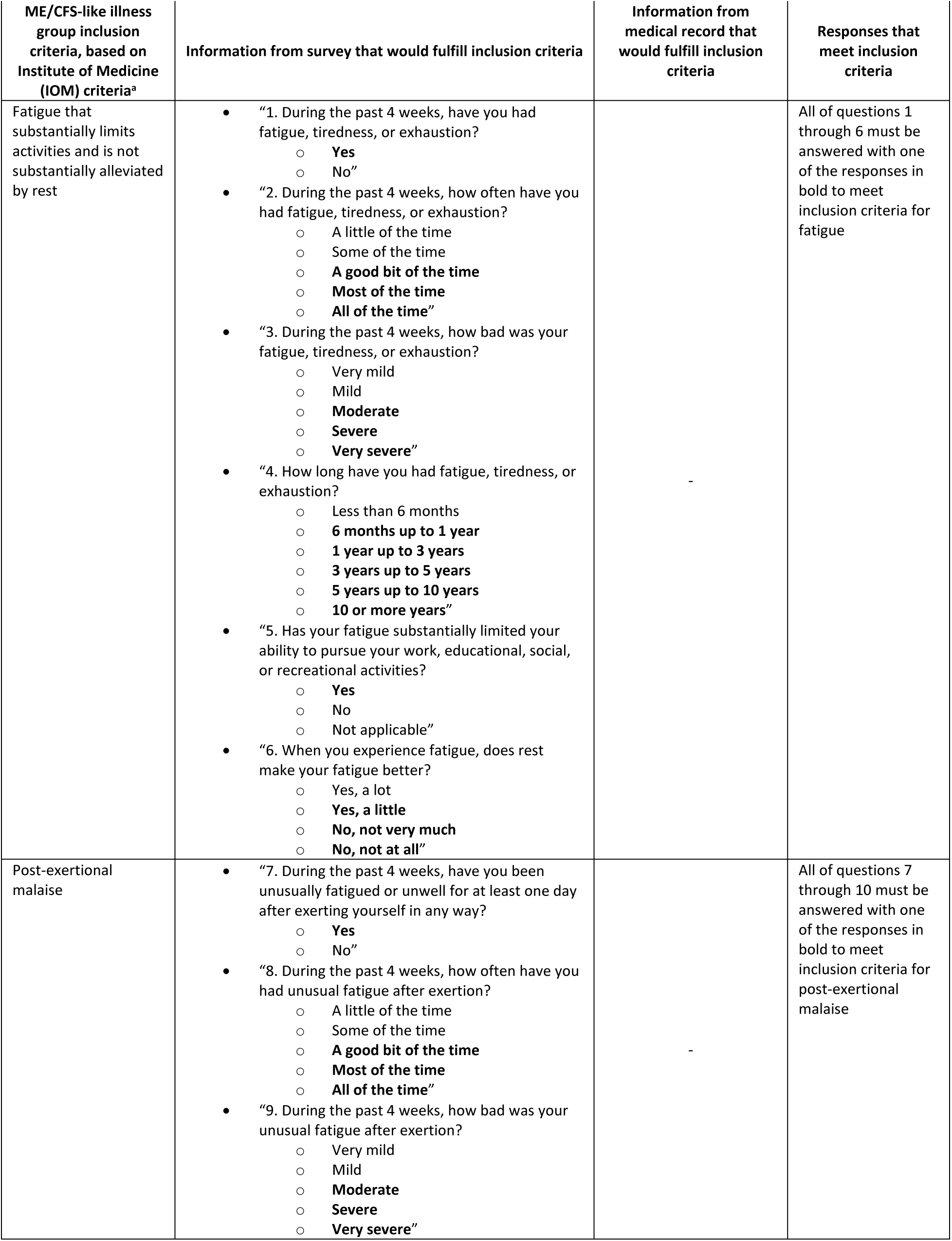
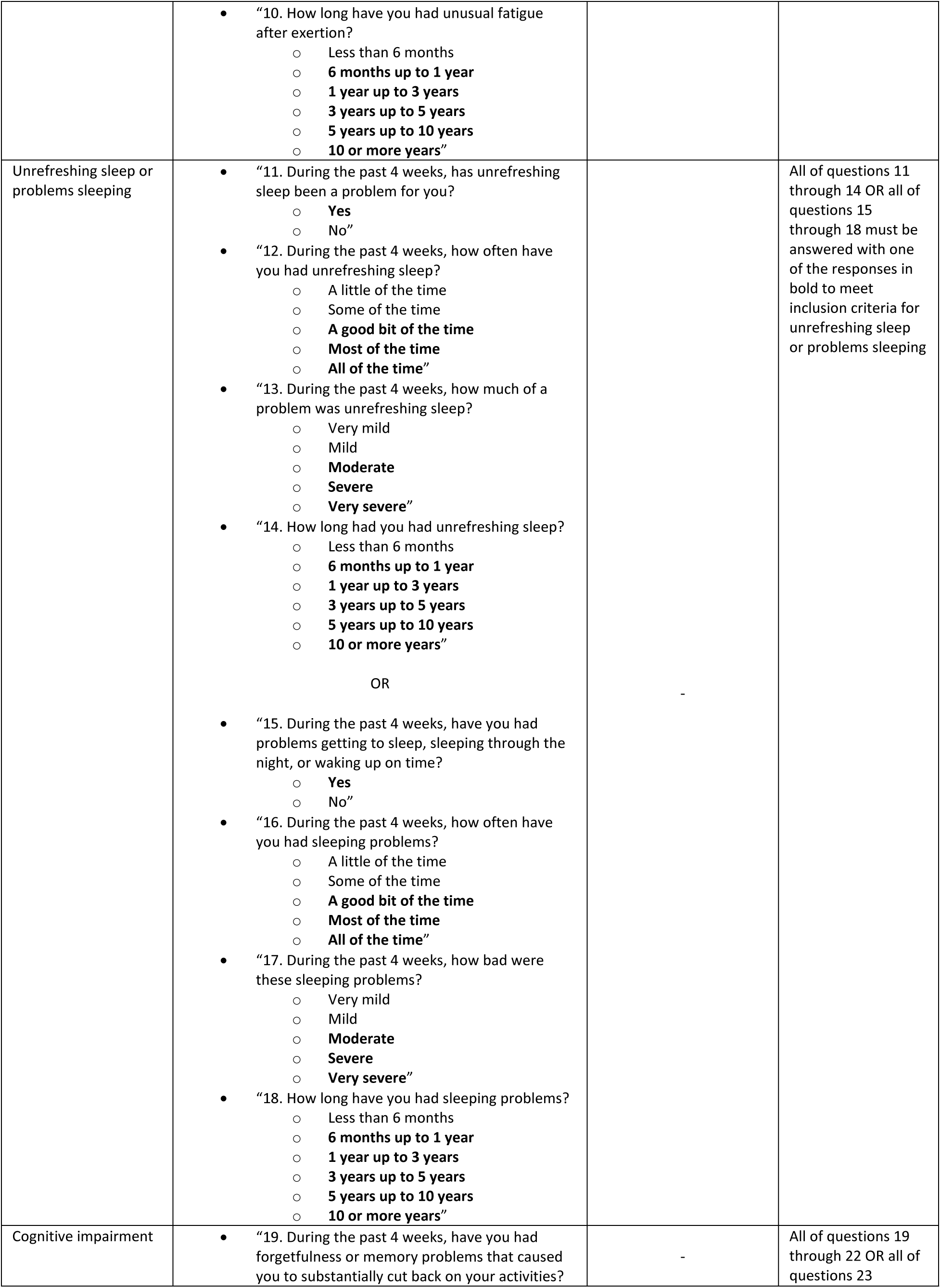
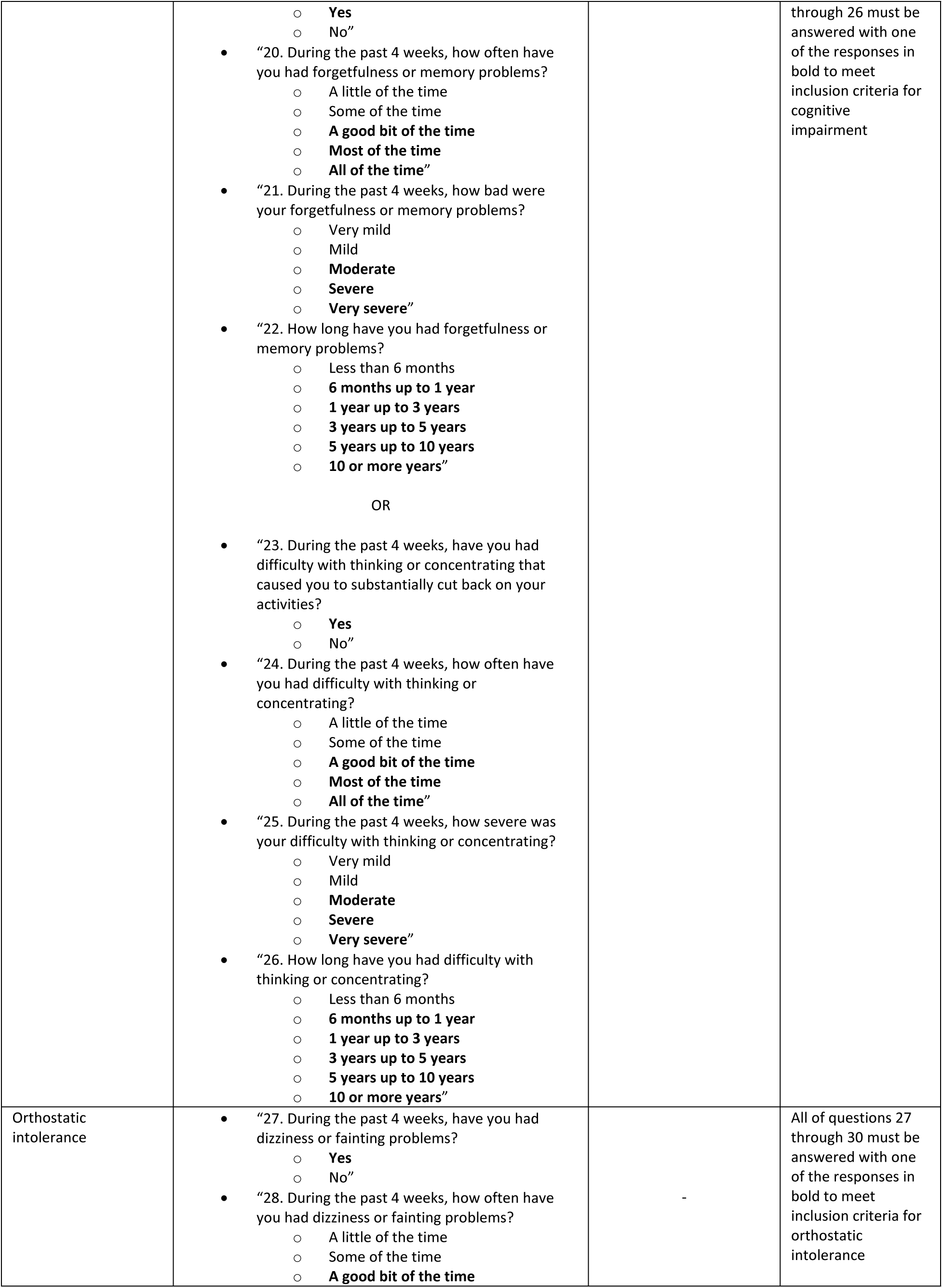
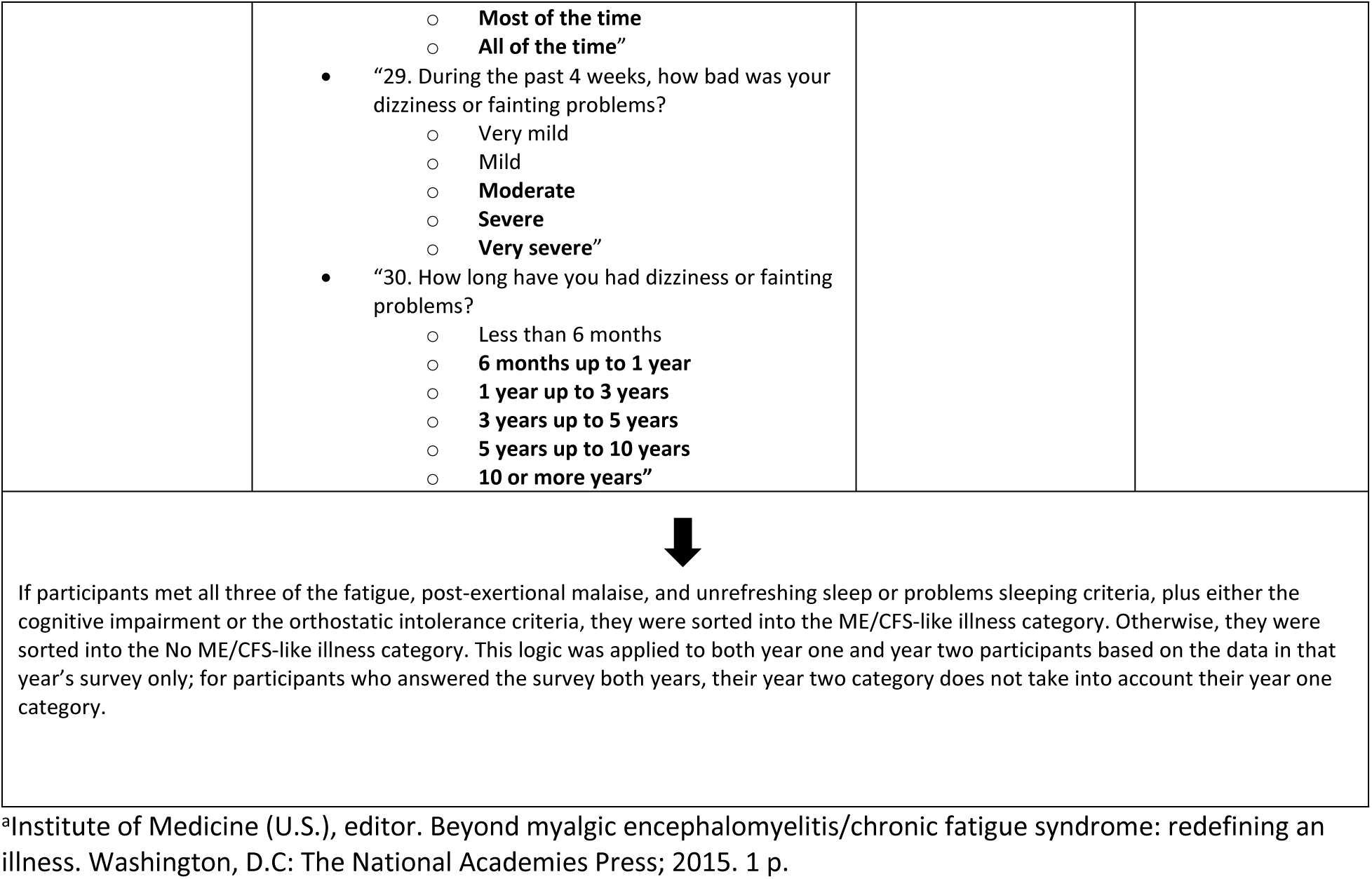
Logic for determining participant grouping into either myalgic encephalomyelitis/chronic fatigue syndrome (ME/CFS)-like illness or No ME/CFS-like illness.

| ME/CFS-like illness group inclusion criteria, based on Institute of Medicine (IOM) criteria<sup>a</sup> | Information from survey that would fulfill inclusion criteria | Information from medical record that would fulfill inclusion criteria | Responses that meet inclusion criteria |
| --- | --- | --- | --- |
| Fatigue that substantially limits activities and is not substantially alleviated by rest | “1. During the past 4 weeks, have you had fatigue, tiredness, or exhaustion?<br>**Yes**<br>No”<br>“2. During the past 4 weeks, how often have you had fatigue, tiredness, or exhaustion?<br>A little of the time<br>Some of the time<br>**A good bit of the time**<br>**Most of the time**<br>**All of the time”**<br>“3. During the past 4 weeks, how bad was your fatigue, tiredness, or exhaustion?<br>Very mild<br>Mild<br>**Moderate**<br>**Severe**<br>**Very severe”**<br>“4. How long have you had fatigue, tiredness, or exhaustion?<br>Less than 6 months<br>**6 months up to 1 year**<br>**1 year up to 3 years**<br>**3 years up to 5 years**<br>**5 years up to 10 years**<br>**10 or more years”**<br>“5. Has your fatigue substantially limited your ability to pursue your work, educational, social, or recreational activities?<br>**Yes**<br>No<br>Not applicable”<br>“6. When you experience fatigue, does rest make your fatigue better?<br>Yes, a lot<br>**Yes, a little**<br>**No, not very much**<br>**No, not at all”** | - | All of questions 1 through 6 must be answered with one of the responses in bold to meet inclusion criteria for fatigue |
| Post-exertional malaise | “7. During the past 4 weeks, have you been unusually fatigued or unwell for at least one day after exerting yourself in any way?<br>**Yes**<br>No”<br>“8. During the past 4 weeks, how often have you had unusual fatigue after exertion?<br>A little of the time<br>Some of the time<br>**A good bit of the time**<br>**Most of the time**<br>**All of the time”**<br>“9. During the past 4 weeks, how bad was your unusual fatigue after exertion?<br>Very mild<br>Mild<br>**Moderate**<br>**Severe**<br>**Very severe”** | - | All of questions 7 through 10 must be answered with one of the responses in bold to meet inclusion criteria for post-exertional malaise |
|  | “10. How long have you had unusual fatigue after exertion?<br>Less than 6 months<br>**6 months up to 1 year**<br>**1 year up to 3 years**<br>**3 years up to 5 years**<br>**5 years up to 10 years**<br>**10 or more years”** |  |  |
| Unrefreshing sleep or problems sleeping | “11. During the past 4 weeks, has unrefreshing sleep been a problem for you?<br>**Yes**<br>No”<br>“12. During the past 4 weeks, how often have you had unrefreshing sleep?<br>A little of the time<br>Some of the time<br>**A good bit of the time**<br>**Most of the time**<br>**All of the time”**<br>“13. During the past 4 weeks, how much of a problem was unrefreshing sleep?<br>Very mild<br>Mild<br>**Moderate**<br>**Severe**<br>**Very severe”**<br>“14. How long had you had unrefreshing sleep?<br>Less than 6 months<br>**6 months up to 1 year**<br>**1 year up to 3 years**<br>**3 years up to 5 years**<br>**5 years up to 10 years**<br>**10 or more years”**<br>OR<br>“15. During the past 4 weeks, have you had problems getting to sleep, sleeping through the night, or waking up on time?<br>**Yes**<br>No”<br>“16. During the past 4 weeks, how often have you had sleeping problems?<br>A little of the time<br>Some of the time<br>**A good bit of the time**<br>**Most of the time**<br>**All of the time”**<br>“17. During the past 4 weeks, how bad were these sleeping problems?<br>Very mild<br>Mild<br>**Moderate**<br>**Severe**<br>**Very severe”**<br>“18. How long have you had sleeping problems?<br>Less than 6 months<br>**6 months up to 1 year**<br>**1 year up to 3 years**<br>**3 years up to 5 years**<br>**5 years up to 10 years**<br>**10 or more years”** | - | All of questions 11 through 14 OR all of questions 15 through 18 must be answered with one of the responses in bold to meet inclusion criteria for unrefreshing sleep or problems sleeping |
| Cognitive impairment | “19. During the past 4 weeks, have you had forgetfulness or memory problems that caused you to substantially cut back on your activities? | - | All of questions 19 through 22 OR all of questions 23 |
|  | ○ **Yes**<br>○ No”<br>• “20. During the past 4 weeks, how often have you had forgetfulness or memory problems?<br>○ A little of the time<br>○ Some of the time<br>○ **A good bit of the time**<br>○ **Most of the time**<br>○ **All of the time”**<br>• “21. During the past 4 weeks, how bad were your forgetfulness or memory problems?<br>○ Very mild<br>○ Mild<br>○ **Moderate**<br>○ **Severe**<br>○ **Very severe”**<br>• “22. How long have you had forgetfulness or memory problems?<br>○ Less than 6 months<br>○ **6 months up to 1 year**<br>○ **1 year up to 3 years**<br>○ **3 years up to 5 years**<br>○ **5 years up to 10 years**<br>○ **10 or more years”**<br>OR<br>• “23. During the past 4 weeks, have you had difficulty with thinking or concentrating that caused you to substantially cut back on your activities?<br>○ **Yes**<br>○ No”<br>• “24. During the past 4 weeks, how often have you had difficulty with thinking or concentrating?<br>○ A little of the time<br>○ Some of the time<br>○ **A good bit of the time**<br>○ **Most of the time**<br>○ **All of the time”**<br>• “25. During the past 4 weeks, how severe was your difficulty with thinking or concentrating?<br>○ Very mild<br>○ Mild<br>○ **Moderate**<br>○ **Severe**<br>○ **Very severe”**<br>• “26. How long have you had difficulty with thinking or concentrating?<br>○ Less than 6 months<br>○ **6 months up to 1 year**<br>○ **1 year up to 3 years**<br>○ **3 years up to 5 years**<br>○ **5 years up to 10 years**<br>○ **10 or more years”** |  | through 26 must be answered with one of the responses in bold to meet inclusion criteria for cognitive impairment |
| Orthostatic intolerance | • “27. During the past 4 weeks, have you had dizziness or fainting problems?<br>○ **Yes**<br>○ No”<br>• “28. During the past 4 weeks, how often have you had dizziness or fainting problems?<br>○ A little of the time<br>○ Some of the time<br>○ **A good bit of the time** | - | All of questions 27 through 30 must be answered with one of the responses in bold to meet inclusion criteria for orthostatic intolerance |
|  | ○ **Most of the time**<br>○ **All of the time**<br>• “29. During the past 4 weeks, how bad was your dizziness or fainting problems?<br>○ Very mild<br>○ Mild<br>○ **Moderate**<br>○ **Severe**<br>○ **Very severe**<br>• “30. How long have you had dizziness or fainting problems?<br>○ Less than 6 months<br>○ **6 months up to 1 year**<br>○ **1 year up to 3 years**<br>○ **3 years up to 5 years**<br>○ **5 years up to 10 years**<br>○ **10 or more years** |  |  |
| If participants met all three of the fatigue, post-exertional malaise, and unrefreshing sleep or problems sleeping criteria, plus either the cognitive impairment or the orthostatic intolerance criteria, they were sorted into the ME/CFS-like illness category. Otherwise, they were sorted into the No ME/CFS-like illness category. This logic was applied to both year one and year two participants based on the data in that year’s survey only; for participants who answered the survey both years, their year two category does not take into account their year one category. |  |  |  |

<sup>a</sup>Institute of Medicine (U.S.), editor. Beyond myalgic encephalomyelitis/chronic fatigue syndrome: redefining an illness. Washington, D.C: The National Academies Press; 2015. 1 p.

